# Neural mechanisms of psychedelic visual imagery

**DOI:** 10.1101/2022.09.07.22279700

**Authors:** Devon Stoliker, Katrin H. Preller, Leonardo Novelli, Alan Anticevic, Gary F. Egan, Franz X. Vollenweider, Adeel Razi

## Abstract

Visual alterations under classic psychedelics can include rich phenomenological accounts of eyes-closed imagery. Preclinical evidence suggests agonism of the 5-HT2A receptor may reduce synaptic gain to produce psychedelic-induced imagery. However, this has not been investigated in humans. To infer the directed connectivity changes to visual sensory connectivity underlying psychedelic visual imagery in healthy adults, a double-blind, randomised, placebo-controlled, cross-over study was performed, and dynamic causal modelling was applied to the resting state eyes-closed functional MRI scans of 24 subjects after administration of 0.2mg/kg of the serotonergic psychedelic drug, psilocybin (magic mushrooms), or placebo. The effective connectivity model included the early visual area, fusiform gyrus, intraparietal sulcus, and inferior frontal gyrus. We observed a pattern of increased self-inhibition of both early visual and higher visual-association regions under psilocybin that was consistent with preclinical findings. We also observed a pattern of reduced inhibition from visual-association regions to earlier visual areas that indicated top-down connectivity is enhanced during visual imagery. The results were associated with behavioural measures taken immediately after the scans, suggesting psilocybin-induced decreased sensitivity to neural inputs is associated with the perception of eyes-closed visual imagery. The findings inform our basic and clinical understanding of visual perception. They reveal neural mechanisms that, by affecting balance, may increase the impact of top-down feedback connectivity on perception, which could contribute to the visual imagery seen with eyes-closed during psychedelic experiences.

## Introduction

Our faculty of imagination allows us to extend perceptual boundaries beyond the immediate sensory world. However, the capacity to visualise imagery is typically distinct from reality during our waking state. When a serotonergic psychedelic is ingested, these restrictions can be altered. This study investigates the neural mechanisms of psychedelic-induced eyes-closed visual imagery (i.e., hallucinations) using psilocybin, a prototypical classic psychedelic derived from select species of mushrooms. Psychedelics allow non-voluntary hallucinated visual imagery to be embedded in an experimental design, which may otherwise be difficult due to the unpredictable occurrence of hallucinations in clinical disorders (1). This research can help us understand the relationship between neuropharmacology and large-scale neural connectivity of the visual system that informs our understanding of perception and clinical hallucinations.

Our investigation into serotonergic psychedelics’ facilitation of visual imagery naturally juxtaposes with conditions like aphantasia, characterised by the inability to voluntarily generate visual imagery (2). This comparison underscores the intricate relationship between neuropharmacology and visual cognition, suggesting that insights from psychedelic research could elucidate the neural basis of visual imagery, the vividness of which varies among individuals (3).

Psychedelic perceptual changes are primarily derived from agonism of the serotonergic 5-HT2A receptor (5-HT2AR) (4–7). The agonism of psychedelic molecules binding to the 5-HT2AR produce an excitatory effect on layer 5 pyramidal neurons which are suggested to modulate neural gain (Burt et al 2021). The 5-HT2AR is highly expressed in the visual and frontal areas of the brain. However, psychedelics may alter neuronal activity in these areas of the brain differently. Under serotonergic psychedelics, frontal area functional connectivity — which describes the temporal synchrony of activity among brain regions— appears to desynchronise (8). Frontal regions serve roles in self-related thinking, planning and egocentric perspective that under psychedelics have been associated with the effect of *ego dissolution* (i.e., decreased self-referential awareness and perception of unity between the self and the world) (9–14). In contrast, posterior visual areas of the brain undergo an opposite transformation and show increased synchrony of functional connectivity that may be associated with change to 5HT2AR (8). For example, evidence for the role of the 5-HT2AR in visual effects (i.e., hallucinations) has been demonstrated in Parkinson’s disease (15).

Psychedelic-induced visual effects can occur with eyes-open or eyes-closed. Review of eyes-closed visual effects suggests they can occur at different levels of complexity and some subjective reports suggest the visualisations can form imagery that match the vividness and realism of eyes-open stimuli (16–19). The most rudimentary forms of eyes-closed visual effects include elementary imagery. Elementary imagery comprises light flashes, reorganising and moving line orientations and geometrical figures containing recurrent patterns (20). These effects have been associated with the early visual area (EVA; corresponding to the primary visual cortex) and may be accompanied by visual intensification such as increased brightness or contrast (19, 21, 22). More phenomenologically rich visual imagery can also occur under psychedelics to induce visualisations of semantic content such as scenes, people, and objects. These visualisations are identified as complex imagery and have been associated with visual and associative connectivity (21–23). Complex imagery is also a form of visual hallucination witnessed in conditions of visual impairment such as Charles Bonnet Syndrome (24, 25). Complex imagery and elemental imagery under psychedelics have been assessed independently using the five dimensions of altered states of consciousness scale, which is a retrospective self-report measure commonly used in psychedelic research (26), see Supplementary for further information ^1^.

Preclinical research investigating change to neural mechanisms of the visual system in the mid 20th-century found that semi-synthetic psychedelic Lysergic diethylamide acid (LSD) decreased neuronal responses of the lateral geniculate nucleus in response to optical tract stimulation of anesthetised cats (27). These findings were supported by contemporary preclinical research that used extracellular recordings and wide-field calcium imaging, which reported that imbalanced 5-HT1A/2A activation reduced sensory drive of the mouse visual cortex (28). Similar research using wide-field two-photon calcium imaging and single-unit electrophysiology of mice under the 5-HT2AR agonist DOI (2,5-dimethoxy-4-iodoamphetamine) also identified reduced net inhibition of visual response amplitude and surround suppression (eyes-open)^2^, while visual feature tuning properties such as retinotopic organization and receptive fields were maintained (30).

The influence of alpha inhibition in the visual system during eyes-closed may also be relevant to psychedelic eyes-closed imagery. Basic preclinical research that measured alpha inhibition found that when our eyes are closed, alpha waves (8-12 Hz range) inhibit activity in the visual system (31, 32). Preclinical research in mice measured single-cell neural firing rates under activation of the 5-HT2AR and identified suppressed high firing rates that are typically indicative of visual stimuli (19, 33–35). Follow-up clinical investigation of alpha inhibition under psilocybin corroborated these results by detecting inverted (i.e., dampened) response of occipital-parietal alpha oscillations to eyes-open and eyes-closed states (36, 37). The reduced alpha inhibition was associated with reduced attention to sensory stimuli during eyes-open and decreased alpha inhibition during eyes-closed periods. This may indicate activity in visual connectivity with eyes-closed that resembles activity typically reserved for eyes-open stimuli (19, 37). Similar findings are associated with eyes-closed visual alterations under the psychedelic N,N-Dimethyltryptamine (DMT), which also identified reduced low frequencies in parietal-occipital regions associated with eyes-closed states (Timmermann et al., 2019). Increased activity in the visual system in the functional connectivity measurements of human subjects under LSD identified the early visual system responds comparably to receiving localised spatial inputs during eyes-closed (38). These functional connectivity changes may have been induced by reduced alpha inhibition of visual connectivity during eyes-closed.

Responses of the visual system to stimulus-triggered perception and internally generated visual imagery rely on similar neural mechanisms (39), which suggest an intricate balance of top-down functional connectivity without the bottom-up processing typical in sensory perception (40). This framework indicates the potential for psychedelics to enhance these mechanisms in eyes-closed states, leading to vivid visualisations. This idea is further reinforced by the evidence that occipital-parietal regions reduce low frequency bands during dreaming (41, 42). However, the primary visual cortex is insufficient for the generation of imagery (43). This underscores the complexity of these neural interactions, highlighting the need for mechanistic analyses to elucidate the distinct yet interconnected connectivity pathways underlying perception and imagery.

To date, the available evidence suggests that psychedelics reduce stimulus-evoked responses along the primary visual pathway during eyes-open conditions. Conversely, during eyes-closed conditions, they appear to disrupt inhibitory mechanisms that control activity in the visual system and increase activity resembling the inputs of external stimuli, potentially indicating that top-down associative inputs influence visual imagery. However, the estimation of sensory inputs in the visual system under psychedelic-induced 5-HT2A receptor agonism have not been investigated in humans. Our study investigated regions of the visual system previously established to be associated with mental imagery (44). The brain regions included the EVA, fusiform gyrus (FG), intraparietal sulcus (IPS) and inferior frontal gyrus (IFG). The EVA is the first stage of cortical visual processing and has demonstrated increased top-down connectivity from visual-association regions to the EVA during eyes-closed visual imagery tasks (44). Connectivity from the EVA follows two broad pathways. The dorsal pathway is connected with the fusiform gyrus (FG), which is part of the inferior temporal cortex that functions to recognise objects and faces (45). The ventral pathway from the EVA is connected to the intraparietal sulcus (IPS), which, along with the inferior frontal gyrus (IFG), is involved in cognitive aspects of imagery. We hypothesised increased self-inhibition and reduced excitatory connectivity of these regions under psilocybin associated with eyes-closed imagery. While our hypothesis of increased self-inhibition and reduced excitatory connectivity under psilocybin diverges from reports of enhanced visual cortex activity in the literature, it is predicated on preclinical findings of decreased synaptic gain under psilocybin facilitating a shift from bottom-up to top-down processing, emphasising internally generated imagery over direct sensory input. This perspective aligns with our focus on eyes-closed conditions, where top-down associative inputs are hypothesised to play a more significant role. We also expected that EVA self-connectivity would be associated with elementary imagery and top-down directed connectivity would be associated with complex imagery.

## Results

We report effective connectivity results in the visual pathways under both placebo and psilocybin conditions and the association of the effectivity with elementary and complex imagery scores obtained after the resting state scan.

Results are reported as the mean effective connectivity of conditions without application of global signal regression (GSR). Multiple design matrices without and with GSR are reported in the Supplementary (see Fig S3, S4 and S5). In Dynamic Causal Modelling (DCM), self-connections are always modelled as inhibitory and log-scaled (see Supplementary for brief technical explanation). Self-connections control the regions’ gain or sensitivity to inputs and the synaptic gain or sensitivity of a region to inputs. A positive self-connection means a relative increased inhibition of the region to external inputs, whereas a negative self-connection means a relative decreased inhibition (i.e., disinhibition) and increased synaptic gain or sensitivity to inputs. Only the self-connections are log-scaled in DCM.

### Visual pathway effective connectivity under placebo

Fig 1, panel (A) displays group-level mean effective connectivity interactions between regions of the visual pathway show inhibition from the IFG to the IPS, FG and EVA. Self-inhibition of the EVA is accompanied by excitation from the FG to the EVA. The results were similar with and without GSR (See Fig S4 and S5 for results with global signal regression).

### Visual pathway effective connectivity change from placebo to psilocybin

Fig 1, panel (B) displays results of the change design matrix, which are calculated from regressors that encode estimations of placebo and the additive effect of psilocybin and are not calculated as the differences between placebo and psilocybin mean effective connectivity (see Supplementary Fig S3 for further details). Our change matrix showed a high degree of coherence with the mean effective connectivity results and demonstrate an alternate complementary view of effective connectivity change estimations induced by psilocybin.

### Visual pathways effective connectivity under psilocybin

Fig 1, panel (C) displays group-level mean effective connectivity between regions of the visual pathway show reduced inhibition from the IFG to the IPS, FG and EVA, as well as between all connections to the EVA, except for the FG to EVA, which remained excitatory. Self-connectivity of all regions is inhibitory, particularly within the EVA and regions of the inferior temporal cortex (FG and IFG) which showed increased inhibitory recurrent connectivity. See Fig S1 for detailed group level region effective connectivity matrix and Table S1 for credible intervals.

**Figure 1.**
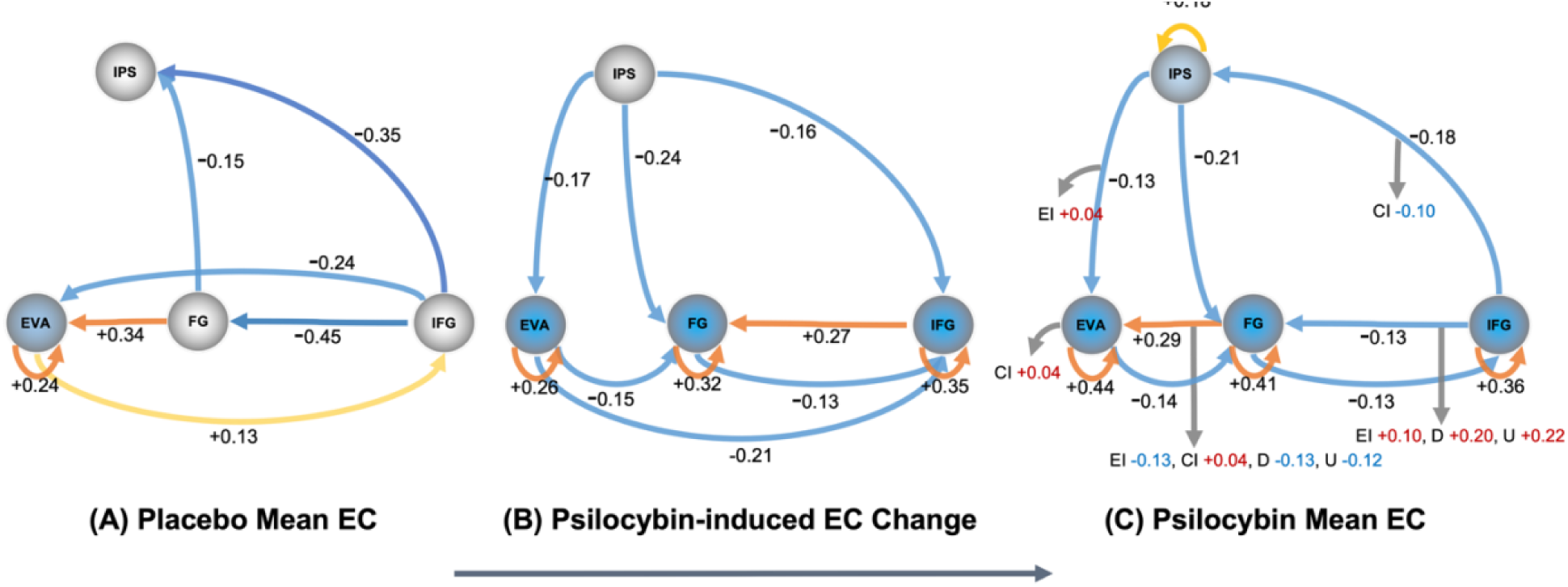
Model of effective connectivity change from placebo to psilocybin. (A) Estimated mean effective connectivity 70 minutes post-administration of placebo, (B) Estimated change to effective connectivity from placebo to psilocybin and (C) Estimated mean effective connectivity 70 minutes post-administration of psilocybin. Brain regions – Early Visual Area (EVA), Fusiform Gyrus (FG), Inferior Frontal Gyrus (IFG), Intraparietal Sulcus (IPS). Cool colours represent inhibition; warm colours represent excitation. For self-connections only, a positive value indicates a move away from excitation, and is represented here as a cool coloured to indicate increased inhibition. Panel (A) displays placebo mean effective connectivity (EC) for reference. Panel (B) shows change from placebo EC which takes into account EC estimates of placebo and psilocybin, 70 minutes post administration to provide posterior expectations (see Supplementary Fig 3 (A) and (B)). These estimations of EC change from placebo to psilocybin reinforce the general trend of mean EC findings and are displayed to aid visualisation of the connectivity changes induced by psilocybin. Unlike mean EC of the group at (A) placebo and (C) under psilocybin, change in posterior expectation is calculated by using the posterior means of EC of both conditions but calculated as the Bayesian model average at the group level taken over the two conditions. Panel (C) displays mean EC changes under psilocybin. Self-inhibition of the EVA, FG, IFG and IPS was found. Panel (B) reaffirmed the EVA, FG and IFG self-inhibition. Excitation from the EVA to IFG was absent in psilocybin mean EC (Panel (C)) and can be explained by the inhibitory change displayed in panel (B). The strong inhibitory connectivity estimated from the IFG to the FG was diminished. Inhibition is demonstrated across all efferent connections except the FG, which remained at similar effective size values (posterior expectations) under placebo and psilocybin. Panel (C) also demonstrates behavioural associations to mean EC, 70 minutes post administration of psilocybin and indicates the direction of EC change found under psilocybin. Behavioural scores were measured on a shortform 5D-ASC in-scanner, immediately after scans. A positive behavioural association, indicated by a red value, means the behavioural variable tends to increase when the EC moves in the excitatory direction. Conversely, a negative association, indicated by a blue value, means the behavioural variable tends to decrease when the EC moves in the excitatory direction. Values for EI = elemental imagery, CI = complex imagery, D = disembodiment and U = experience of unity are displayed. The effect sizes (i.e., the posterior expectations) of connections are in Hz except self-connections which are modeled as always inhibitory and are log-scaled (see Supplementary material for more details). Behavioural associations and connections displayed are estimated at posterior probability > 0.99, which amounts to very strong evidence. For results inclusive of posterior probability > 0.50, see Supplementary Fig S2.

### Behavioural associations to effective connectivity under psilocybin

Fig 1, panel (C) displays behavioural scores acquired in-scanner, immediately after the scans measured elemental imagery and complex imagery. Associations between group-level mean effective connectivity and elemental and complex imagery were analysed. Non-visual subjective effects (disembodiment and experience of unity) were also collected in-scanner and were also analysed. Connectivity change towards excitation, identified in the connectivity from the IFG to the FG under psilocybin, was positively associated with elemental imagery, disembodiment, and an experience of unity. Connectivity from the FG to EVA were associated with all subjective effects, however, this connection did not show strong evidence of change from placebo to psilocybin. All other associations showed increased inhibition of the connection associated with an increase in the subjective effects. See Fig 1 and Fig S2 for the respective connectivity matrices.

## Discussion

When we open our eyes, visual perception appears to be driven by the stimuli of the outside world. However, consciousness research has identified the significant role of the brain in constructing visual perception. The German physicist and physician, Hermann von Helmholtz said, “Objects are always imagined as being present in the field of vision as would have to be there in order to produce the same impression on the nervous mechanism”. This axiom suggests visual perception of the external world manifests from the inference of neural activity. Neural activity corresponding to vision is guided by signals from the sensory epithelia of the eyes. However, the eyes’ ability to represent the environment is limited by foveal acuity, subcortical filtration mechanisms and attention processes. Prior knowledge, recognition and context, such as visual schemas, help determine the likely causes of sensory sensations and signify the importance of beliefs and associations in the construction of visual perception. For example, inferences in visual perception rely on expectations. Expectations that influence perception may be learned, for example, prior expectations have been shown to facilitate hallucinations (46) or prior expectations may be innate, demonstrated by visual illusions and perceptual biases, such as the Adelson’s checker-shadow illusion (47). These examples and previous research demonstrate that visual perception is configured by intrinsic neural mechanisms and influenced by high-level cognitive inferences.

Our investigation measured the large-scale expression of psychedelic neuropharmacological changes on brain connectivity to give insights into the neural mechanisms that underlie the perception of eyes-closed visual imagery in human subjects under psilocybin. The findings revealed a pattern of inhibition in the effective connectivity of self-connections of visual pathways of subjects under psilocybin. DCM estimated that inhibition in the self-connections of all regions in our model under the influence of psilocybin was greater than the inhibition seen in subjects under placebo (see Figs 1 and 2). The results of the current study align with previous preclinical findings that indicate that reduced sensory drive enhances internal transmissions under agonism of the 5-HT2AR (as reported by Azimi et al., 2020; Evarts et al., 1955; Michaiel et al., 2019). The reduced sensory drive appears to generalise to the effective connectivity of the human visual system under psilocybin and suggests the augmentation of internal signals may induce visual imagery, in the absence of eyes-open sensory stimuli. The increased self-inhibition of visual and associative regions indicates a decrease in synaptic gain that corresponds to the decreased sensitivity of these regions to inputs. Additionally, it is noteworthy to mention that psilocybin reduced sensitivity to external stimuli under eyes-open conditions (48). The decreased synaptic gain may relate to previous findings of reduced parieto-occipital alpha oscillations measured in the 8-12Hz band under psilocybin, which may facilitate spontaneous activation in eyes-closed states (as reported by Michael Kometer et al., 2013; M. Kometer & Vollenweider, 2018). Although the decreased synaptic gain appears to contradict increased activity in the visual cortex and the synchronised temporal patterns identified in sensory regions by functional connectivity analysis (as reported by Roseman et al., 2016; Katrin H. Preller et al., 2020), these discrepancies may originate from measurement techniques and analysis methods.

Visual effects related to the visual functions of EVA, FG and IFG may be associated with the self-inhibition of these regions. The FG is strongly involved in the processing and recognition of objects and faces (45), while the EVA receives, segments, and integrates visual information (47). We found top-down connectivity from the IFG to FG and FG to EVA associated with elementary imagery and complex imagery, and damage to this connectivity has previously been associated with Charles Bonnet syndrome hallucinations (49). However, evidence between effective connectivity changes and their visual effects converged for only the IFG to FG and FG to EVA connections. Moreover, the association between these connection and subjective effects was not exclusive to visual effects scores only and were also statistically associated with disembodiment and experience of unity. Although the roles of EVA, FG and IFG connectivity in visual literature reinforces the likelihood of their involvement in visual effects, the connectivity’s association to non-visual effects indicate the behavioural associations we report are preliminary. See Supplementary Fig S2 and limitations below for details.

Feedforward (bottom-up) excitatory effective connectivity from the EVA to IFG under psilocybin showed inhibition in our model of change. Previous fMRI research of healthy adults has estimated the effective connectivity when voluntarily imagining objects (44). This research indicated group activity from the IFG to the EVA was more excitatory when objects were imagined versus when visually observed, suggesting that sensory perception is underwritten by feedforward connectivity while imagery is underwritten by feedback (top-down) connectivity. Our findings show the inhibition if the feedforward connection, suggesting psilocybin’s effect on connectivity between the IFG and EVA move in the same direction as seen in effective connectivity during voluntarily imagined objects, showing a trend of reduced feedforward and increased feedback connectivity. The differences between our findings and that of previous voluntarily imaginary research which use the same model of regions may mark the contrast between task-based imagery that requires intentional imagination and our experimental design which measured spontaneous pharmacologically-induced imagery.

The IFG, which has a role in cognitive processes during visual working memory (50), showed greater top-down connectivity under psilocybin that is demonstrated by its efferent connections (see Fig 1). For example, we found top-down inhibitory connectivity from IFG to IPS was reduced compared to placebo and was associated with complex imagery. The IPS is involved in visual attention and the maintenance and manipulation of spatial information in working memory (51). The reduced top-down inhibition under psilocybin may suggest top-down inferences are amplified and serve a role in imagery.

An alternative model of psychedelic-altered consciousness, the RElaxed Beliefs Under pSychedelics (REBUS model) (8, 52), suggests reduced top-down (feedback) connectivity and increased bottom-up (feedforward) connectivity underlies the neural mechanisms of psychedelics. While this model may apply to alternate bottom-up connectivity to specific cortical connections (see (53)), our findings do not support the bottom-up principle of this model between visual and associative regions. Instead, we found a pattern of reduced inhibitory top-down effective connectivity and strong self-inhibition of visual and associative regions. Psilocybin-induced effects in our model are interpreted to sensitise visual connectivity to top-down endogenous signals. Greater reliance on top-down priors that we have identified is aligned with a strong priors model of the visual system that has previously been used to describe the occurrence of hallucinations (46) and highly relevant related lines of enquiry exploring strengthened priors underlying psychedelic visual effects (see (54)). Schizophrenia research which investigated different pharmacological mechanisms, also described how impaired sensory input may allow attentional mechanisms a preponderant role that leads to hallucinations (55).

Therefore, our findings suggest reduced sensory drive may amplify the role of top-down inferences involved in visual perception in a manner similar to dreaming. To support this perspective we point to the reduced parieto-occipital alpha oscillations detected both in dreams and under psychedelics with eyes-closed (37, 41, 54). Alpha waves, which are suggested to inhibit brain regions not undergoing operations, disappear during sleep, however, their role during dreams remains ambiguous (56–58). Relatedly, the experience of sensory synaesthesias, such as visual imagery driven by music, is common under psychedelics (59) and suggests the delineation of signals may be diminished under psychedelics. The novel interaction of signals may allow top-down associative inferences to manifest as visual imagery.

Future connectivity research could measure interactions between visual, emotional and association connectivity to investigate the mechanisms that control the content and complexity of psychedelic imagery. This pursuit may also advance therapeutic applications using psychedelics (9). For example, research has described the contents of complex visual imagery can take on a personalised meaning that may support therapeutic changes (60). Psychedelics may also be tested on populations diagnosed with aphantasia, who typically cannot perceive visual imagery, to explore the differential impact of psychedelics on connectivity between the frontal and posterior regions involved in visual imagery production (2, 61). This could help reveal distinct neural mechanisms at play in aphantasia.

Future research studies could also measure effective connectivity changes between visual and associative regions at higher psychedelic doses to advance our understanding of the integrative function between sensory and associative connectivity. For example, it has been suggested that the difficulty to differentiate whether the source of a signal represents reality or imagination, known as reality monitoring, depends upon top-down connectivity to earlier visual areas, such as the IFG to the EVA (62). Higher doses of psilocybin which result in experiences of *ego dissolution* and related out-of-body experiences can give insight into the mechanisms that underlie these symptoms in clinical disorders (10). For example, higher doses would provide more reasons to examine reality monitoring; however, in this study, participants received only a low to moderate dose. We found the IFG self-connectivity, which serves a role in cognitive processes during visual working memory (50), was diminished (inhibited) under psilocybin. The effective connectivity between the IFG to EVA may be an important mechanism for reality monitoring that future research could investigate at increasing doses of psychedelics.

Our investigation of imagery in a healthy population under psilocybin has several limitations. A notable limitation of our study derives from the strong correlation among various psychedelic effects induced by psilocybin. This co-occurrence of subjective effects, which range from elementary and complex imagery to feelings of disembodiment and experiences of unity, indicate intrinsic associations among self-reported subjective experiences, which in turn produce correlated scores on in-scanner rating scales. This poses challenges in the precise statistical separation between subjective effects and their association with connectivity patterns, and the distinct link of visual effects to connectivity within our analytical framework. For example, our model identified connectivity associated with the non-visual subjective experiences of disembodiment and experience of unity. Our brain and behavioural associations should therefore be treated with caution as a preliminary evidence of statistical regularities rather than definitive causal links between neural connectivity and visual experiences (see Supplementary for further discussion). We contrast the behaviour-connectivity limitations by pointing to the models’ regions established role in visual processing, its use in previous visual imagery research (see (44)) and the strength we demonstrate in the consistency of our model’s effective connectivity results, with and without the use of global signal regression, and across different design matrices (see Supplementary Fig S3, S4 and S5). Other limitations include the selection of brain regions and their coordinates of location, which may influence the results. In addition to replication using the same regions, future research may explore an alternate selection of regions, including established visual circuits, and alternate methods to determine their coordinates. Imagery tasks may also facilitate group-specific activations and the identification of coordinates in the regions of interest. For discussion of regions that warrant further investigation, such as the thalamus, and regions involved in visual sense of space, see Supplementary discussion. Imaging that accommodates subjective differences in the onset of visual imagery is also needed. Furthermore, the psychedelic dose can strongly impact the results. Participants were given less than a standard clinical dose of psilocybin typically used during therapeutic interventions (i.e., 25 mg). The vividness of imagery and connectivity dynamics may be more altered at a higher dose. The small healthy adult sample (n=20 after thresholding for head motion) is also a limitation. Lastly, preclinical research varied between anaesthetised and eyes-open recordings, suggesting the alignment of the present results with preclinical literature requires validation from further clinical studies.

## Conclusion

Closing our eyes typically shields us from the visual sense of the outside world. Psychedelics can penetrate the visual barrier of our eyelids, generating visual imagery in the absence of sensory stimuli. In this research, we explored possible mechanisms underlying the generation of psilocybin-induced visual perception. We demonstrated inhibition of visual brain region inputs and reduced effective connectivity between regions involved in visual processes that occur under psilocybin. The findings suggest that emergence of visual imagery without external visual inputs may be facilitated by reduced sensitivity of visual regions, and inhibition of visual connectivity in our model, indicative of a hierarchical shift in balance towards feedback signaling. Our results extend previous preclinical research that found 5-HT2AR inhibits the stimulus-evoked activity of visual regions and add to the existing knowledge of clinical hallucinations and neural mechanisms of visual imagery. Future investigations with varied dosages, brain regions, and task-based designs will further our understanding of the complex effects of psychedelics on visual perception.

By altering perception, psychedelics provide a means of measuring changes in the mechanisms responsible for perceptual interpretation of the world. The ability of psychedelics to induce visual alterations and imagery expands the scope for perception and clinical research alongside the investigation of psychiatric disorders, brain injury, sleep and dreams. The presence of imagery in eyes-closed states indicates that psychedelics impact the brain’s reliance on sensory input for distinguishing between subjective experiences and objective reality. This aligns with the broader concept of psychedelics dissolving the barriers between the subject and object, highlighting their profound effect on our understanding of consciousness and perception.

## Methods

### Participants

The data analysed in this paper were collected as part of a previous study (registered at ClinicalTrials.gov (NCT03736980)), which is reported in (6) and was approved by the Cantonal Ethics Committee of Zurich. 24 subjects (12 males and 11 females; mean age = 26.30 years; range = 20–40 years) were recruited through advertisements at universities in Zurich, Switzerland. The Mini-International Neuropsychiatric Interview (MINI-SCID) (63), the DSM-IV fourth edition self-rating questionnaire for Axis-II personality disorders (SCID-II) (64), and the Hopkins Symptom Checklist (SCL-90-R) (65) were used to exclude subjects with present or previous psychiatric disorders or a history of major psychiatric disorders in first-degree relatives.

Sample size was determined based on a previous study reporting psychedelic-induced effects on functional brain connectivity (66). See Supplementary for further detail.

### Design

The data we analyse is part of a double-blind, randomised, placebo-controlled, cross-over study was performed once. At 2 different occasions at minimum 2 weeks apart, each participant received either Placebo (179-mg mannitol and colloidal silicon dioxide [Aerosil; Evonik Resource Efficiency GmbH] 1-mg orally or Psilocybin (0.2-mg/kg body weight, orally). Resting state scans (10 minutes each) were taken 20-, 40- and 70-minutes following administration of psilocybin or placebo. However, only scans at 70 minutes during the peak effects were used in our analysis. Participants were asked to not engage in repetitive thoughts such as counting. Furthermore, participants were asked to close their eyes during the resting state scan. A short version of the 5 Dimensions of Altered States of Consciousness Scale (5D-ASC, a retrospective self-report questionnaire) (26, 67) that included 4 subdimensions (elementary imagery, complex imagery, disembodiment and experience of unity) was administered in-scanner immediately after the scan to assess the time course of subjective effects. The questions were displayed with MR-compatible video goggles Resonance Technology Inc., Northridge, USA). Participants answered the questions on a scale ranging from 1 (not at all) to 4 (very much) using a 4-button response box.

### MRI Data Acquisition and Preprocessing

MRI data were acquired on a Philips Achieva 3.0T whole-body scanner. A 32-channel receive head coil and MultiTransmit parallel radio frequency transmission was used. 265 volumes were acquired per resting state scan resulting in a scan duration of 10 mins. See Supplementary for more details. The acquired images were analysed using SPM12 (68). The pre-processing steps of the images consisted of slice-timing correction, realignment, spatial normalization to the standard EPI template of the Montreal Neurological Institute (MNI), and spatial smoothing using a Gaussian kernel of 6-mm full-width at half maximum. Head motion was investigated for any excessive movement. 3 subjects were excluded due to excessive head motion and one subject did not complete the scan at 70 minutes so eventually 20 subjects entered our connectivity analysis pipeline.

### Extraction of Region Coordinates Across Subjects

Regions, model complexity and ROI size were selected based on prior literature (44, 69–71). A generalized linear model (GLM) was used to regress 6 head motion parameters (3 translation and 3 rotational), white matter and cerebrospinal fluid signals from preprocessed data. Global signal regression (GSR) was not used in our pre-processing pipeline to align with visual research of the 2A receptor (28, 30). However, the results with GSR are reported in Supplementary to provide consistency with line with our previous investigations (14). The time series for each ROI was computed as the first principal component of the voxels activity within an 8 mm sphere centred on the region of interest (ROI) coordinates listed in Table 1.

**Table 1.** Regions of Interest and MNI coordinates.

| <u>Region</u> | <u>MNI coordinates</u> |  |  |
| --- | --- | --- | --- |
|  | *x* | *y* | *z* |
| Early visual area (EVA) | 28 | -94 | 4 |
| Fusiform gyrus (FG) | 33 | -60 | -13 |
| Intraparietal sulcus (IPS) | 33 | -59 | 53 |
| Inferior frontal gyrus (IFG) | 46 | 17 | 2 |

**Figure 2.**
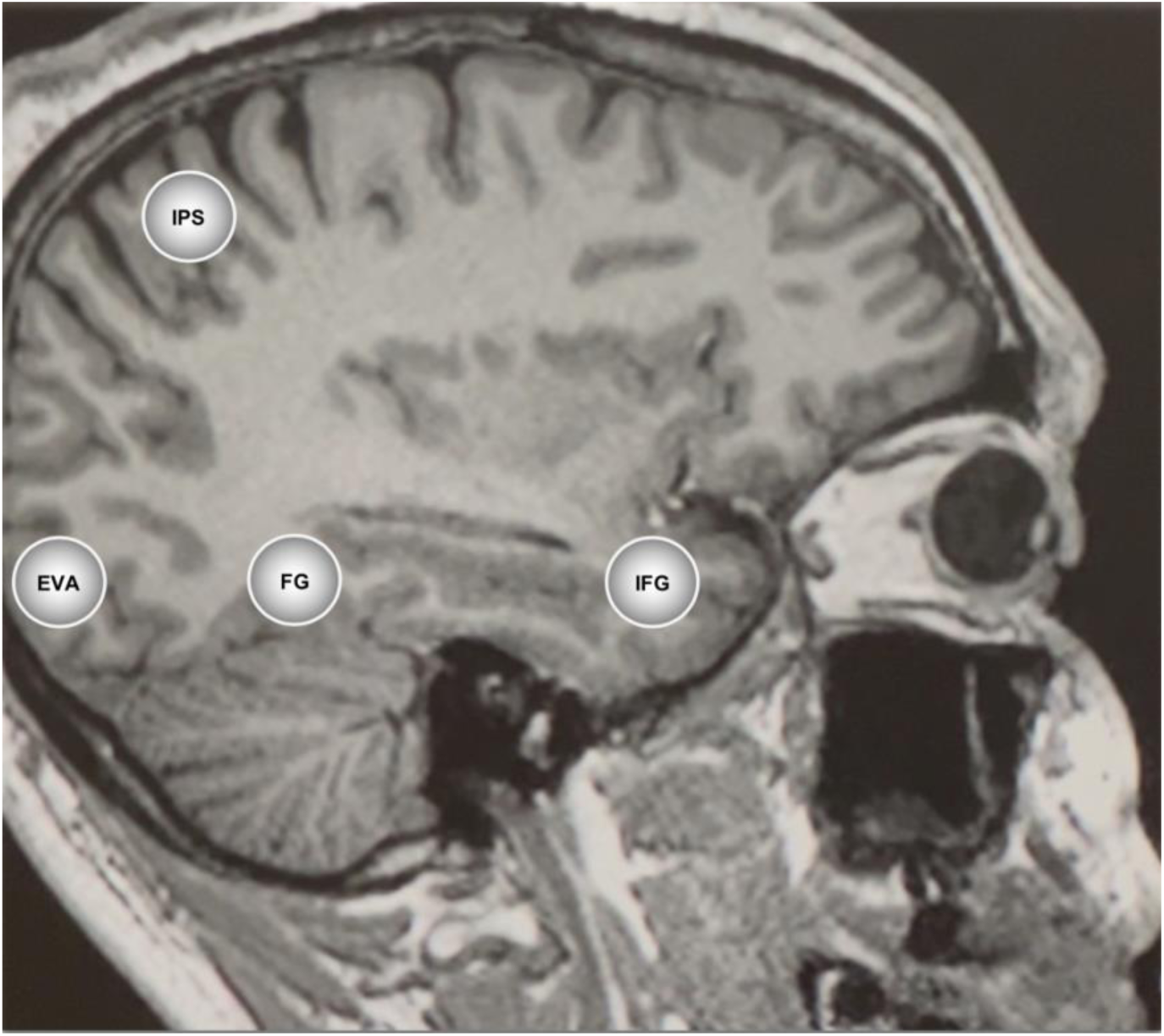
Coordinates of regions of interest. The visual and associative regions comprised of the early visual cortex (EVC), fusiform gyrus (FG); inferior parietal sulcus (IPS) and inferior frontal gyrus (IFG).

### Specification and Inversion of DCM

Dynamic causal modelling (DCM) enables measurement of the direction of connectivity among circuits, which relies on Bayesian model comparison demonstrated to discriminate between inhibition and excitation of bottom-up (feedforward) and top-down (feedback) connections (72, 73). This analysis technique allows non-invasive inference of the directed connectivity and self-connectivity (i.e., synaptic activity).

A fully-connected DCM model was specified using the four ROIs identified in Table 1, without any exogenous inputs. The DCM for each subject was then inverted using spectral DCM (73, 74) to infer the effective connectivity that best explains the observed cross-spectral density for each subject. The DCM fit to the data using cross spectral density averaged 87.3% for placebo conditions and 86.3% for psilocybin conditions.

### Second Level Analysis Using Parametric Empirical Bayes

The effective connectivity inferred by spectral DCM for each subject is taken to the second (group) level to test hypotheses about between-subject effects. A Bayesian General Linear Model (GLM) is employed to characterise individual differences in effective connectivity into hypothesised group-average connection strengths plus any unexplained noise. Hypotheses on the group-level parameters are tested within the Parametric Empirical Bayes (PEB) framework (75), where both the expected values and the covariance of the connectivity parameters are taken into account. That is, precise parameter estimates influence the group-level result more strongly than uncertain estimates, which are down-weighted. Bayesian model reduction is used as an efficient method of Bayesian model selection (75).

In DCM, effective connectivity is measured in the unit of Hertz (Hz). The unit of Hz quantifies the rate of change in (neural states) per unit time that one brain region causes in another. Unit of Hz refer to directed connections. Self-connections are log-scaled, hence have no unit.

### Behavioural Associations

Parametric Empirical Bayes (PEB) was used to estimate the effective connectivity associations with subjective scores. The posterior probabilities express the likeliness of association between neural connectivity estimates and behavioural scores. We applied a statistical threshold of posterior probability > 0.99 (equivalent to a Bayes Factor of > 150 and considered a “very strong” evidence (76) to estimate the significance of meaningful relationships between observed neural patterns and measured behavioural outcomes. See Supplementary for further technical details about PEB and Discussion for limitations of behavioural associations.

## Data Availability

All data produced in the present study are available upon reasonable request to the authors

## Acknowledgments and Disclosures

All data are available in the main text, Supplementary, or by request to the corresponding author. The authors report no biomedical financial interests or potential conflicts of interest.

## Footnotes

1 See Kometer and Vollenweider 2018 for a review of Serotonin Hallucinogen-Induced Visual Perceptual Alterations 19. Kometer M, Vollenweider FX. Serotonergic Hallucinogen-Induced Visual Perceptual Alterations. Curr Top Behav Neurosci. 2018;36:257-82..

2 Surround suppression refers to the presence of neighbouring stimuli diminishing the neural response to a central visual stimulus in the primary visual cortex 29. Schallmo MP, Murray SO. Identifying separate components of surround suppression. J Vis. 2016;16(1):2..

