## Supplementary Materials for "Neural mechanisms of psychedelic visual imagery"

Devon Stoliker, Monash Biomedical Imaging, 762-772 Blackburn Rd, Clayton VIC 3168. Australia.

+ Joint Senior Authors

### Spectral Dynamic Causal Modelling

Dynamic causal modelling (DCM) is Bayesian framework that infers the directed (causal) connectivity among the neuronal systems – referred to as effective connectivity. We recently proposed a new DCM for resting state fMRI – based upon a deterministic model that generates predicted cross spectra – referred to as spectral DCM. In order to model resting state activity – in the absence of external stimuli – we will have to add a stochastic component, i.e. neural fluctuations, to the classical DCM based on ordinary differential equations. Mathematically, we can express the formulation of the stochastic generative model using a set of two equations. First is the neuronal state equation, namely

$$\dot{x}(t) = f(x(t), u(t), \theta) + v(t), \tag{S1}$$

and second is the observation equation, which is a static nonlinear mapping from the hidden physiological states in (1) to the observed BOLD activity and is written as:

$$y(t) = h(x(t), \varphi) + e(t), \tag{S2}$$

where $\dot{x}(\mathrm{t})$ is the rate of change of the neuronal states $x(t)$, $\theta$ are unknown parameters (i.e. the effective connectivity) and $v(t)$ (resp. $e(t)$) is the stochastic process – called the state noise (resp. the measurement or observation noise) – modelling the random neuronal fluctuations that drive the resting state activity. In the observation equations, $\varphi$ are the unknown parameters of the (haemodynamic) observation function and $u(t)$ represents any exogenous (or experimental) inputs that drive the hidden states – that are usually absent in resting state designs (1). Spectral DCM furnishes a constrained inversion of the stochastic model by parameterising the neuronal fluctuations $v(t)$. Spectral DCM simplifies the generative model by replacing the original timeseries with their second-order statistics (i.e., cross spectra). This means, instead of estimating time varying hidden states, we are estimating their covariance which is time invariant. Then we simply need to estimate the covariance of the random fluctuations; where a scale free (power law) form for the state noise (resp. observation noise) is used – motivated from previous work on neuronal activity (2-4) – as follows:

$$g_v(\omega, \theta) = \alpha_v \omega^{-\beta_v}$$

$$g_e(\omega, \theta) = \alpha_e \omega^{-\beta_e} \tag{S3}$$

Here, $\{\alpha, \beta\} \subset \theta$ are the parameters controlling the amplitudes and exponents of the spectral density of the neural fluctuations. The parameterisation of endogenous fluctuations means that the states are no longer probabilistic; hence the inversion scheme is significantly simpler, requiring estimation of only the parameters (and hyperparameters) of the model.

We used standard Bayesian model inversion to infer the parameters of the model in (1), (2) and (3), from the observed signal $y(t)$. The description of the Bayesian model inversion procedures based on variational Laplace can be found elsewhere for the interested readers (5-7).

### Parametric Empirical Bayes

Empirical Bayes refers to the Bayesian inversion or fitting of hierarchical models. In hierarchical models, constraints on the posterior density over model parameters at any given level are provided by the level above. These constraints are called empirical priors because they are informed by empirical data. We recently introduced a second-level or between-subjects model over parameters, which represents how individual (within-subject) connections derive from the subjects' group membership (8) – based on parametric empirical Bayes (PEB). This approach calls on Bayesian Model Reduction (BMR) to finesse the inversion of multiple models of a single dataset or a single (hierarchical) model of multiple datasets. BMR allows one to compute posterior densities over model parameters, under new prior densities, without explicitly inverting the model again. For example, one can invert a DCM for each subject in a group and then evaluate the posterior density over group effects, using the posterior densities over parameters from the single subject inversion. This may improve subject-specific parameter estimates, by using group-level estimates to rescue individual DCM from local optima. Mathematically, for DCM studies with *N* subjects and *M* parameters per DCM, we have a hierarchical model, where the responses of the *i*-th subject and the distribution of the parameters over subjects can be modeled as:

$$y_i = \Gamma_i^{(1)}(\theta^{(1)}) + \varepsilon_i^{(1)} \qquad \text{(S4)}$$

$$\theta^{(1)} = \Gamma^{(2)}\left(\theta^{(2)}\right) + \varepsilon^{(2)}$$

$$\theta^{(2)} = \eta + \varepsilon^{(3)}$$

where, $y_i$ is the BOLD time series from *i-th* subject *and* $\Gamma_i^{(1)}$ is a nonlinear mapping from the parameters of a model to the predicted response $y$ for e.g. as shown in Eq. S1 above. $\varepsilon_i^{(1)}$ is independent and identically distributed (i.i.d.) observation noise (equivalent to $e(t)$ in Eq. S2). In this hierarchical form, *empirical priors* encoding second (between-subject) level effects place constraints on subject-specific parameters. The second level would be a linear model where the random effects are parameterised in terms of their precision:

$$\Gamma^{(2)}\left(\theta^{(2)}\right) = (X \otimes W)\beta$$

where, $\beta \subset \theta$ are group means or effects encoded by a design matrix with between $X$ and within-subject $W$ parts. The between-subject part encodes differences among subjects or covariates such as age, while the within-subject part specifies mixtures of parameters that show random effects. We assume that the first column of the design matrix is a constant term, modelling group means and subsequent columns encode group differences or covariates such as age.

### Self-connections in DCM

Please note that in DCM, the self-connections are always modelled as inhibitory (to preclude any run-away excitation), but these parameters in the model are log-scaled for the sake of numerical stability of the model fitting procedures. This (log) scaling means that these self-connections can take both positive (red) and negative values (blue). A positive self-connection means a relative increased inhibition, whereas a negative self-connection means a relative decreased inhibition (i.e., disinhibition). Inhibitory self-connections control the regions' gain or sensitivity to inputs. Only the self-connections are log-scaled in DCM.

### Subjective Effects (5D-ASC)

Elemental imagery and complex imagery were measured on the retrospective 5D-ASC 70 minutes after the administration of psilocybin and scored between 1-4. Under 0.2mg/kg psilocybin group level elementary imagery averaged = 2.45/4; SD = 1.07; range = 1–4 and complex imagery averaged = 2.80/4; SD = 0.98; range = 1.33–4. Under placebo group level elementary imagery averaged 1.31/4; SD = 0.52; range = 1–3 and complex imagery averaged = 1.60/4; SD = 0.82; range = 1–4.

A long version of the 5D-ASC was also completed by the participants 360 min after drug treatment and scored between 0-100. Elementary imagery averaged = 54.96/100; SD = 31.31; range = 0–97.67 and complex imagery averaged = 51.62/100; SD = 34.71; range = 1–100. Under placebo group level elementary imagery averaged 3.65/100; SD = 7.01; range = 0/00-24.33 and complex imagery averaged = 4.10/100; SD = 6.71; range = 0–26.67.

### Participants

All participants were deemed healthy after screening for medical history, physical examination, blood analysis, and electrocardiography. Participants were asked to abstain from prescription and illicit drug use two weeks prior to first testing and throughout the duration of the study and abstain from alcohol use 24 hours prior to testing days. Urine tests and self-report questionnaires were used to verify the absence of drug and alcohol use. Urine tests were also used to exclude pregnancy. Further exclusion criteria included left-handedness, poor knowledge of the German language, cardiovascular disease, history of head injury or neurological disorder, history of alcohol or illicit drug dependence, MRI exclusion criteria, including claustrophobia, and previous significant adverse reactions to a hallucinogenic drug. All participants provided written informed consent statements in accordance with the declaration of Helsinki before participation in the study. Subjects received written and oral descriptions of the study procedures, as well as details regarding the effects and possible risks of drug treatment.

### MRI Data Acquisition and Preprocessing Additional Details

Images were acquired using a whole-brain gradient-echo planar imaging (EPI) sequence (repetition time, 2,430 ms; echo time, 27 ms; slice thickness, 3 mm; 45 axial slices; no slice gap; field of view, 240 × 240 $mm^2$; in-plane resolution, 3 × 3 $mm^2$; sensitivity-encoding reduction factor, 2.0). Additionally, two high-resolution anatomical images (voxel size, 0.7 × 0.7 × 0.7 $mm^3$) were acquired using T1-weighted and T2-weighted sequences using 3D magnetization prepared rapid-acquisition gradient echo sequence (MP-RAGE) and a turbo spin-echo sequence, respectively. T1-weigthed images were collected via a 3D magnetization-prepared rapid gradient-echo sequence (MP-RAGE) with the following parameters: voxel size= 0.7x0.7x0.7 mm3, time between two inversion pulses= 3123 ms, inversion time= 1055 ms, interecho delay= 12 ms, flip angle= 8°, matrix= 320x335, field of view= 224x235 mm2, 236 sagittal slices. Furthermore T2-weighted images were collected using via a turbo spin-echo sequence with the following parameters: voxel size= 0.7x0.7x0.7 mm3, repetition time= 2500 ms, echo time= 415 ms, flip angle= 90°, matrix= 320x335, field of view= 224x235 mm2, 236 sagittal slices.

### Discussion Addendum

#### Brain and Behavioural Associations

The co-occurrence of subjective effects, which range from elementary and complex imagery to feelings of disembodiment and experiences of unity, show intrinsic associations among self-reported subjective experiences, which in turn produce correlated scores on in-scanner rating scales. This poses challenges in the precise statistical separation between subjective effects and their association with connectivity patterns, and the distinct link of visual effects to connectivity within our analytical framework.

Additional limitations include the behaviour score distribution on our in-scanner measures of the 5D-ASC. Participants responded to imagery measures using a button box that constrained group scores to a 1-4 scale. The limited granularity of the score distribution may affect the analysis of behavioural associations between connectivity and imagery, although alignment between the time of imaging and behavioural measurement ensured the altered visual experience occurred during the scan.

Brain and behavioural associations may be improved by task-based paradigms that use stimuli that selectively amplify the subjective experience of visual phenomena and employing sample sizes sufficiently large to enable the analysis of subsets of participants based on variability in their subjective effects.

#### Future Investigation of Effective Connectivity

Signals that influence visual imagery may derive from connectivity across various circuits throughout the brain. For example, limbic connectivity such as between the retrosplenial cortex, anterior hippocampus, parahippocampus and entorhinal cortex that share roles in egocentric and allocentric perspectives and spatial aspects of imagery (9-12). The cuneus, precuneus, posterior parietal, lingual, and lateral occipital gyrus are also visual regions that serve important functions, often distinguished by the dorsal and ventral visual pathways, that were not investigated in this analysis. Moreover, subcortical regions that have been indicated in models of the action of psychedelics on the brain, such as the thalamus may be valuable for future research in visual effects (13).

**Figure S1.**

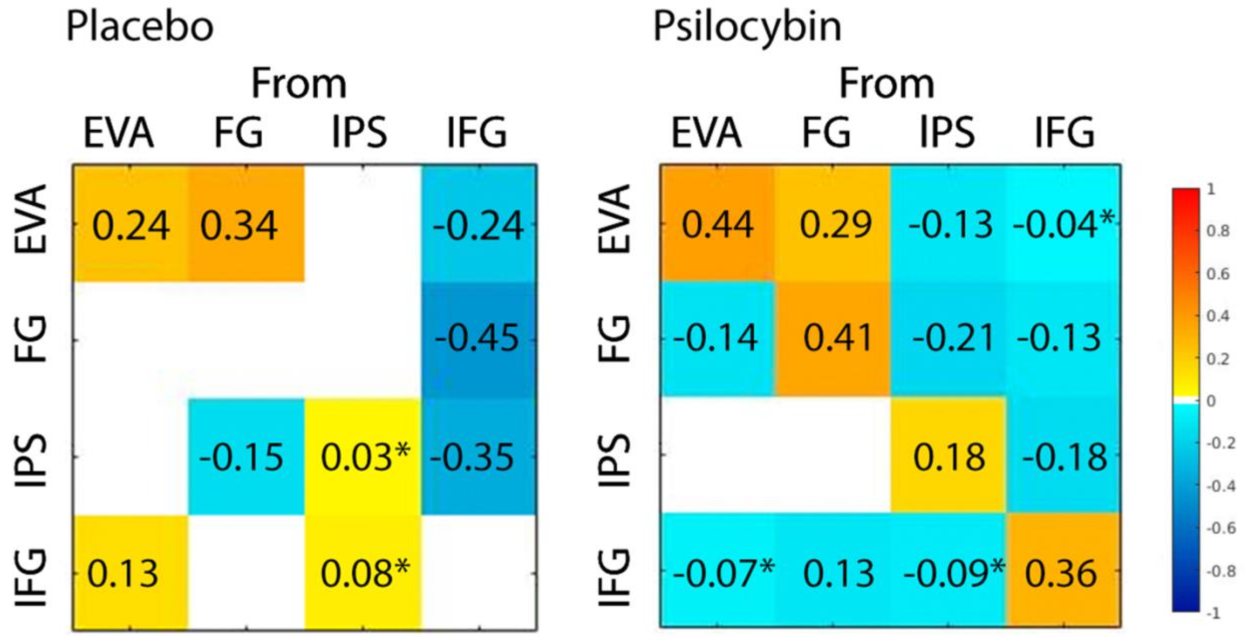


**Group-level region effective connectivity.** Left panel shows placebo matrix. Right panel shows 70 min post psilocybin administration matrix. Values are posterior expectations measured in Hz. Warm coloured connections represent excitatory mean effective connectivity posterior expectations; Cool coloured connections represent inhibitory mean effective connectivity posterior expectations. Please note, increased inhibition of self-connections, here displayed in red, are demonstrated in cool colours in the main manuscript to facilitate interpretation. *Denotes posterior probability threshold > .50. All other values are posterior probability threshold > .99. .99 results correspond to figures in the main manuscript.

**Figure S2.**

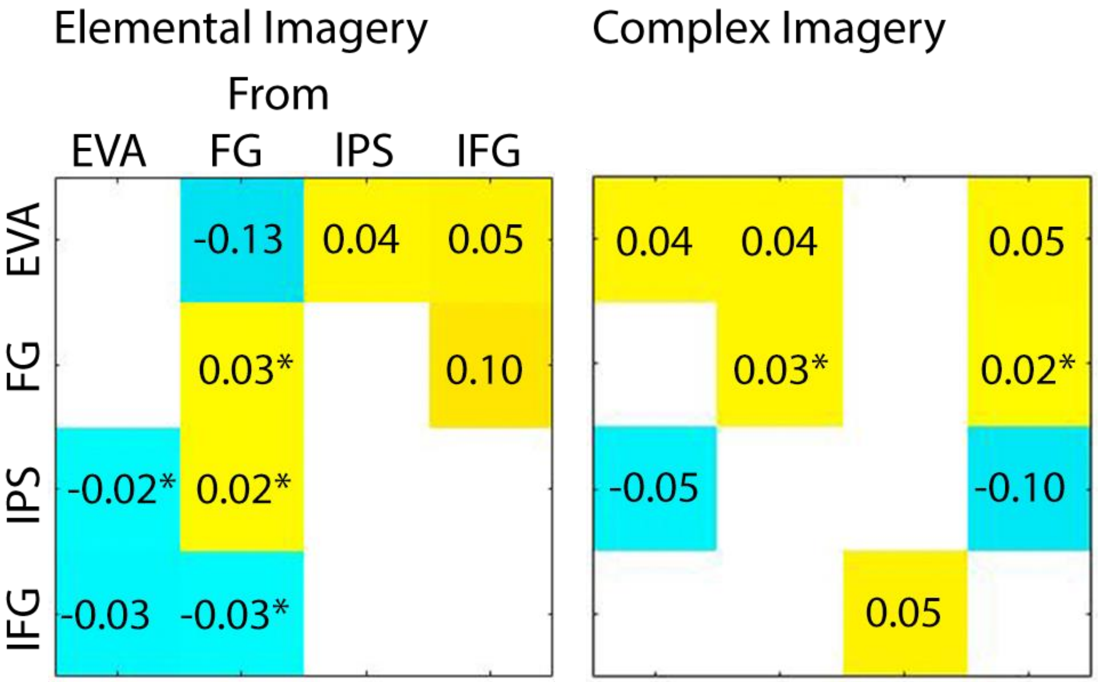


**Associations of group-level region effective connectivity to elementary imagery and complex imagery.** Scores were measured on the 5D-ASC at the end of the scan. Warm colours represent positive associations between directed connection and imagery, cool colours represent negative association between directed connection and imagery. Left panel shows region effective connectivity associations to elemental imagery 70 minutes post psilocybin. Right panel shows region effective connectivity associations to complex imagery 70 minutes post psilocybin. *Denotes posterior probability threshold > .50. All other values are posterior probability threshold > .99.

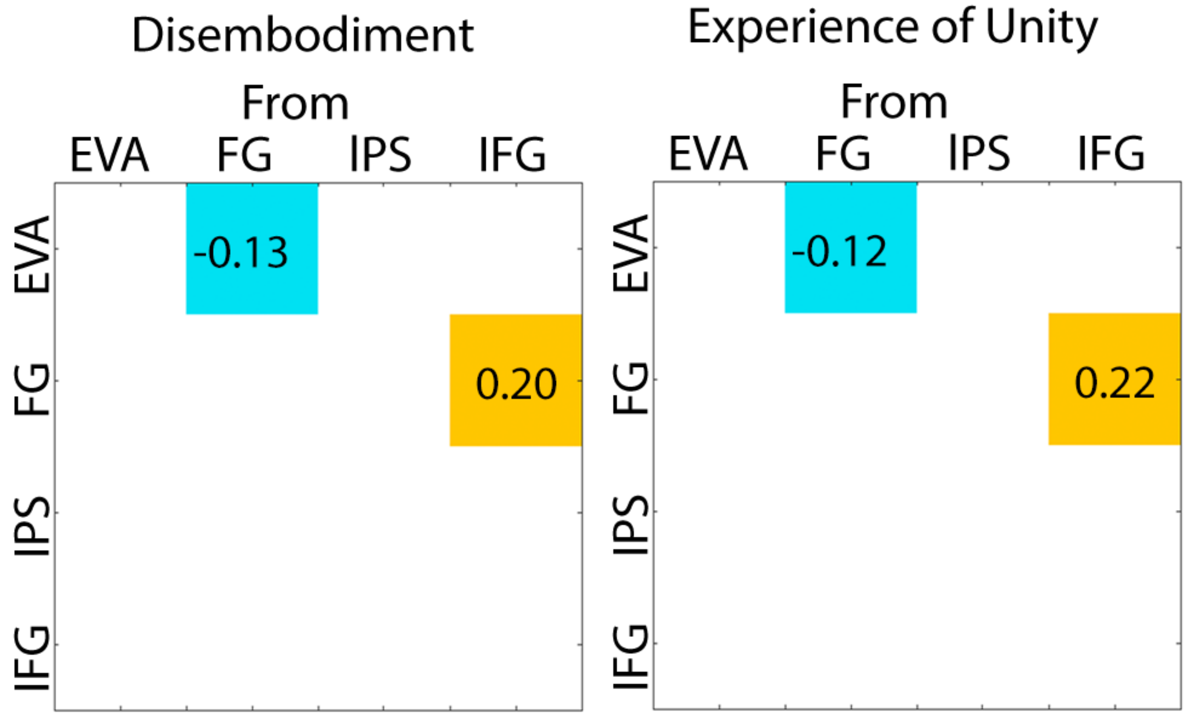


**Associations of group-level region effective connectivity to disembodiment and experience of unity.** Scores were measured on the 5D-ASC at the end of the scan. Warm colours represent positive associations between directed connection and subjective effect, cool colours represent negative association between directed connection and subjective effect. Left panel shows region effective connectivity associations to disembodiment 70 minutes post psilocybin. Right panel shows region effective connectivity associations to experience of unity 70 minutes post psilocybin. Results are shown with both posterior probability threshold > .50 and posterior probability threshold > .99 .

**Figure S3.**

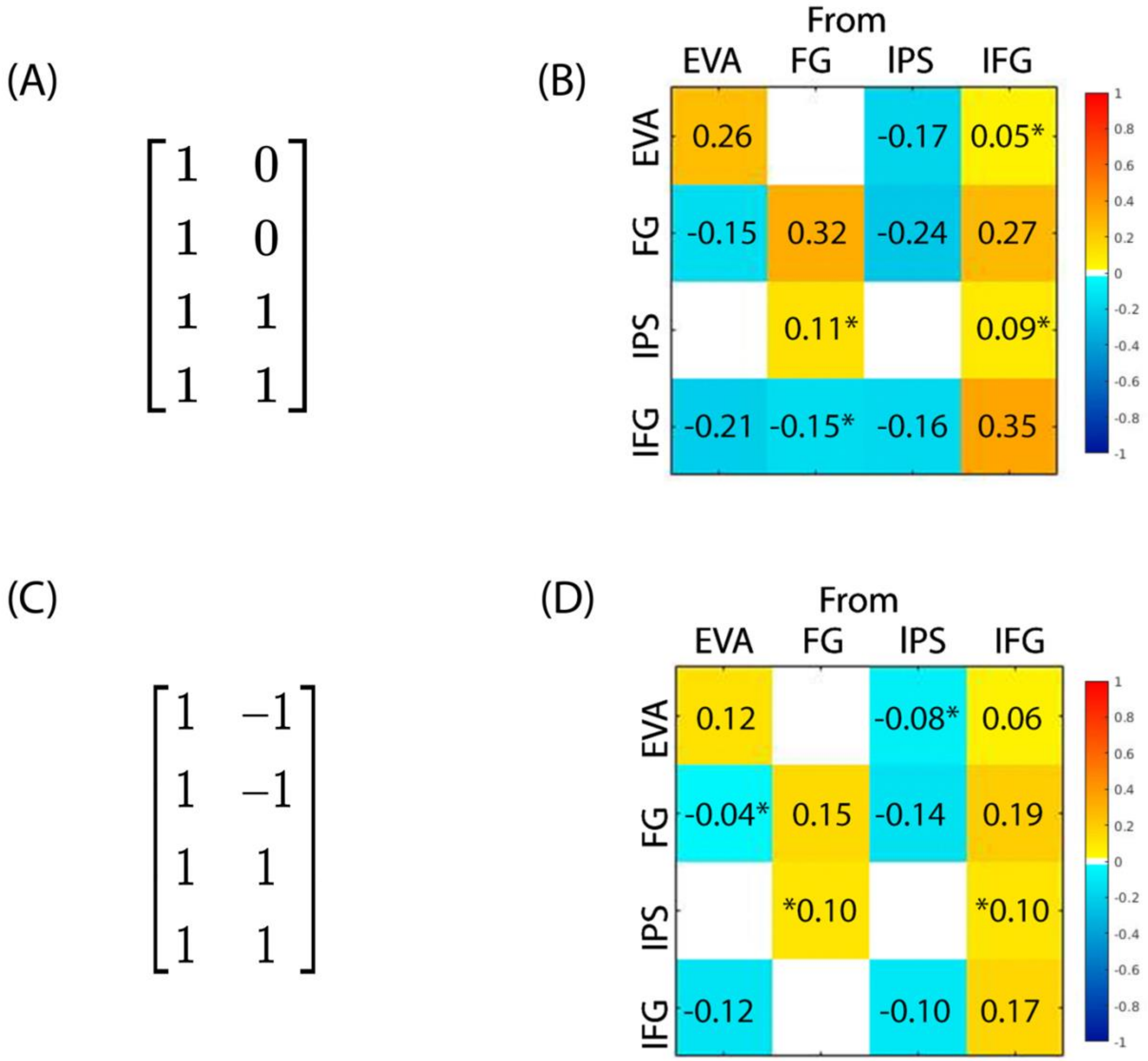


**Alternative design matrix.** Respective design matrix and effective connectivity posterior expectation matrices are demonstrated. (A) Designated the placebo group to serve as the baseline. Regressors in change design matrix encode: 1) placebo group 2) the additive effect of being in the second group (psilocybin after 70 min) relative to the placebo group. See (B) for results. (C) Regressors in the contrast design matrix encode: 1) the group mean and 2) group difference relative to the mean. See (D) for results. Warm coloured connections represent an effective connectivity change towards excitation; Cool coloured connections represent an effective connectivity change towards inhibition. Please note, to facilitate interpretation, increased inhibition of self-connections, here displayed in red, are demonstrated in cool colours in the main manuscript. Values are posterior expectations measured in Hz. *Denotes posterior probability threshold > .50. All other values are posterior probability threshold > .99.

**Figure S4**

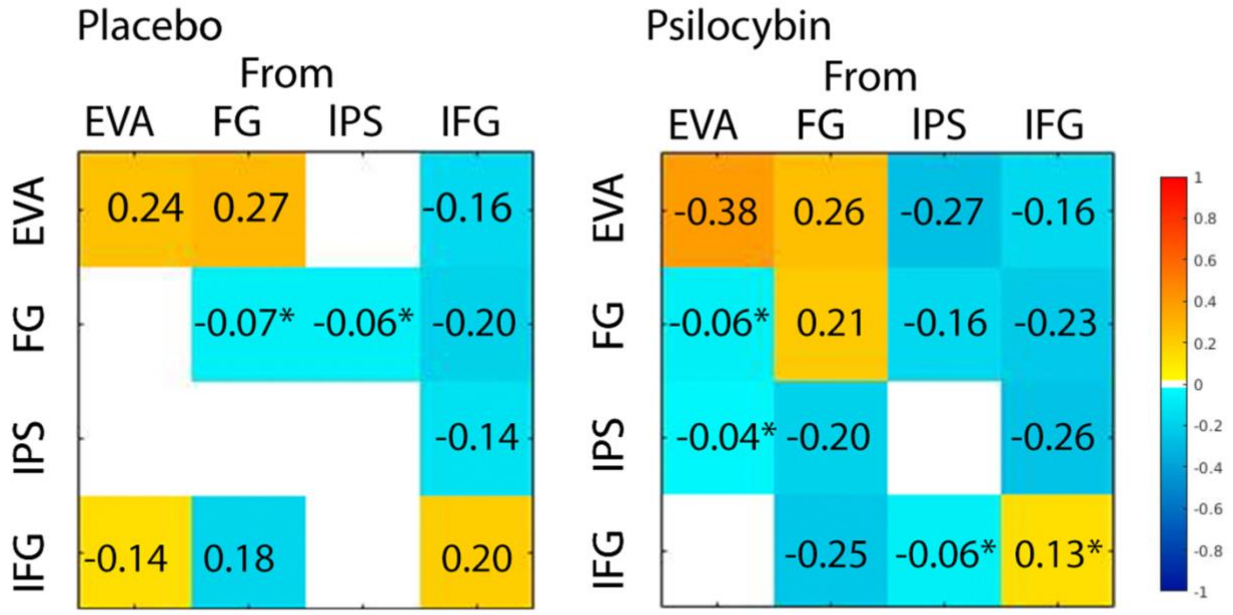


**Group-level region effective connectivity with global signal regression.** (A) placebo, (B) 70 min and post psilocybin administration. Warm coloured connections represent excitatory mean effective connectivity posterior expectations; Cool coloured connections represent inhibitory mean effective connectivity posterior expectations. Please note, increased inhibition of self-connections, here displayed in red, are demonstrated in cool colours in the main manuscript to facilitate interpretation. Values are posterior expectations measured in Hz. *Denotes posterior probability threshold > .50. All other values are posterior probability threshold > .99.

**Figure S5.**

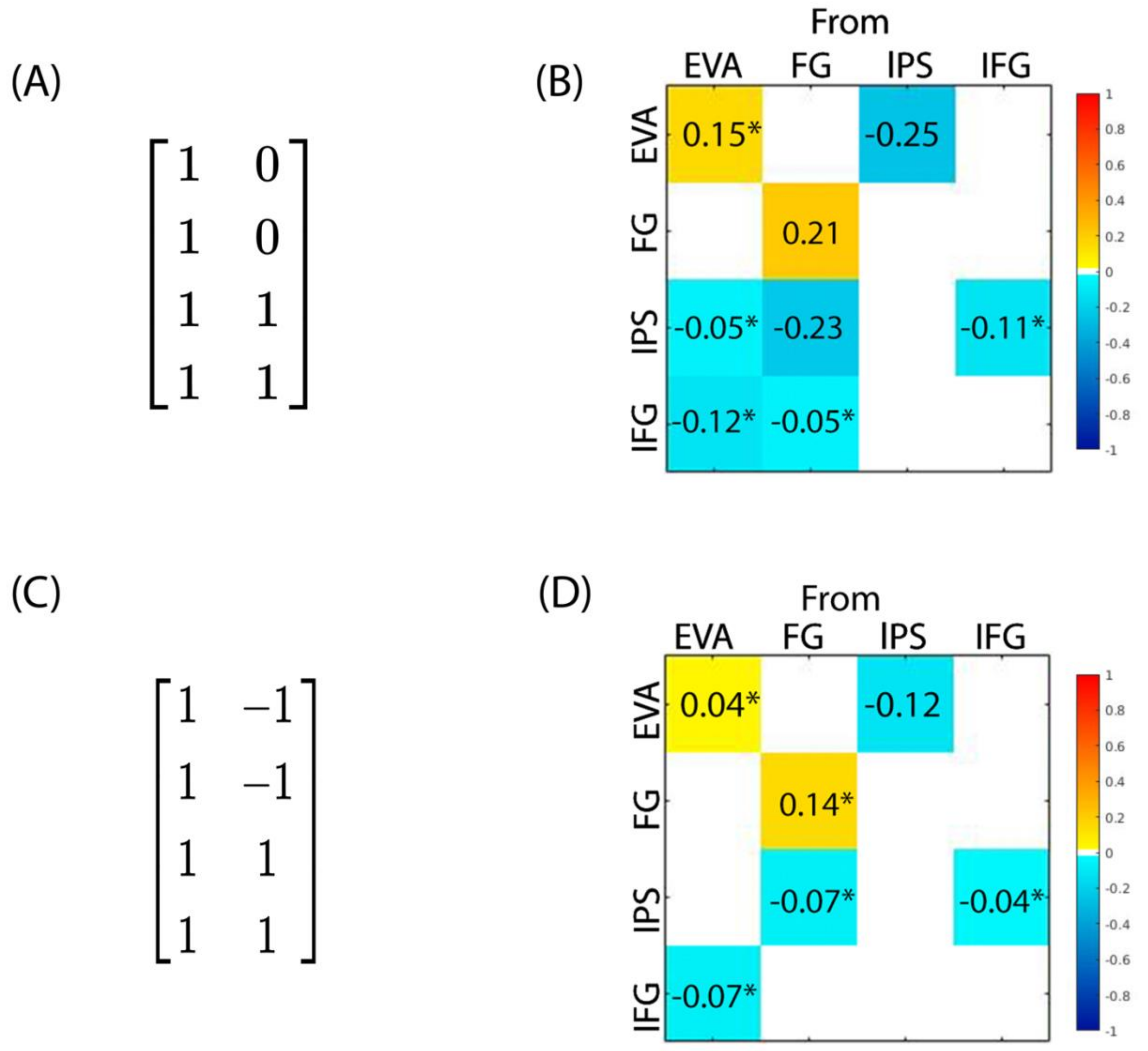


**Alternative design matrix and group-level region effective connectivity with global signal regression.** Respective design matrix and effective connectivity posterior expectation matrices are demonstrated with global signal regression applied. (A) Designated the placebo group to serve as the baseline. Regressors in change design matrix encode: 1) placebo group 2) the additive effect of being in the second group (psilocybin after 70 min) relative to the placebo group. See (B) for results. (C) Regressors in the contrast design matrix encode: 1) the group mean and 2) group difference relative to the mean. See (D) for results. Warm coloured connections represent an effective connectivity change towards excitation; Cool coloured connections represent an effective connectivity change towards inhibition. Please note, to facilitate interpretation, increased inhibition of self-connections, here displayed in red, are demonstrated in cool colours in the main manuscript. Values are posterior expectations measured in Hz. *Denotes posterior probability threshold > .50. All other values are posterior probability threshold > .99.

**Table S1.**

*Placebo*

| *Connection* | *Valence & Effect Size* | *Credible Intervals (low/high)* |
|---|---|---|
| EVA → EVA | +0.24 | 0.183/0.303 |
| FG → EVA | +0.34 | 0.277/0.369 |
| IFG → EVA | -0.24 | -0.305/-0.175 |
| IFG → FG | -0.45 | -0.526/-0.377 |
| FG → IPS | -0.15 | -0.204/-0.090 |
| IFG → IPS | -0.35 | -0.402/-0.296 |
| EVA → IFG | -0.13 | 0.084/0.172 |

*Psilocybin*

| *Connection* | *Valence & Effect Size* | *Credible Intervals (low/high)* |
|---|---|---|
| EVA → EVA | +0.44 | 0.371/0.507 |
| FG → EVA | +0.29 | 0.247/0.331 |
| IPS → EVA | -0.13 | -0.186/-0.080 |
| EVA → FG | -0.14 | -0.196/-0.091 |
| FG → FG | +0.41 | 0.341/0.482 |
| IPS → FG | -0.21 | -0.283/-0.135 |
| IFG → FG | -0.13 | -0.190/-0.063 |
| IPS → IPS | +0.18 | 0.118/0.274 |
| IFG → IPS | -0.18 | -0.249/-0.113 |
| FG → IFG | -0.13 | -0.188/-0.079 |
| IFG → IFG | +0.36 | 0.275/0.437 |

**Mean region effective connectivity.** All results are for posterior probability > 0.99.

*References*


1. Friston KJ, Kahan J, Biswal B, Razi A. A DCM for resting state fMRI. NeuroImage. 2014;94:396-407.
2. Beggs JM, Plenz D. Neuronal avalanches in neocortical circuits. J Neurosci. 2003;23(35):11167-77.
3. Shin CW, Kim S. Self-organized criticality and scale-free properties in emergent functional neural networks. Phys Rev E Stat Nonlin Soft Matter Phys. 2006;74(4 Pt 2):045101.
4. Stam CJ, de Bruin EA. Scale-free dynamics of global functional connectivity in the human brain. Hum Brain Mapp. 2004;22(2):97-109.
5. Razi A, Friston K. The Connected Brain: Causality, models, and intrinsic dynamics. IEEE Signal Processing Magazine. 2016;33(3):14-35.
6. Friston KJ, Harrison L, Penny W. Dynamic causal modelling. Neuroimage. 2003;19(4):1273-302.
7. Friston K, Mattout J, Trujillo-Barreto N, Ashburner J, Penny W. Variational free energy and the Laplace approximation. Neuroimage. 2007;34(1):220-34.
8. Friston KJ, Litvak V, Oswal A, Razi A, Stephan KE, van Wijk BC, et al. Bayesian model reduction and empirical Bayes for group (DCM) studies. Neuroimage. 2016;128:413-31.
9. Alexander Andrew S, Carstensen Lucas C, Hinman James R, Raudies F, Chapman GW, Hasselmo Michael E. Egocentric boundary vector tuning of the retrosplenial cortex. Science Advances. 2020;6(8):eaaz2322.
10. Kim M, Maguire EA. Hippocampus, retrosplenial and parahippocampal cortices encode multi-compartment 3D space in a hierarchical manner. bioRxiv. 2018:200576.
11. Vann SD, Aggleton JP, Maguire EA. What does the retrosplenial cortex do? Nature Reviews Neuroscience. 2009;10(11):792-802.
12. Doeller Christian F, Kaplan R. Parahippocampal Cortex: Translating Vision into Space. Current Biology. 2011;21(15):R589-R91.
13. Preller KH, Razi A, Zeidman P, Stämpfli P, Friston KJ, Vollenweider FX. Effective connectivity changes in LSD-induced altered states of consciousness in humans. Proceedings of the National Academy of Sciences. 2019;116(7):2743-8.
